# Lifetime vs 10-year Cardiovascular Disease Prediction in Young Adults Using Statistical Machine Learning and Deep Learning: The CARDIA Study

**DOI:** 10.1101/2022.09.22.22280254

**Authors:** Bharath Ambale-Venkatesh, Hieu T. Nguyen, Jared P. Reis, Colin O. Wu, Jeffrey J. Carr, Chike Nwabuo, Samuel S. Gidding, Eliseo Guallar, João A.C. Lima

## Abstract

**Importance:** National guidelines for primary prevention suggest consideration of lifetime risk for cardiovascular (CV) disease in addition to 10-year risk, however, it is unclear if the predictors of 10-year vs lifetime (10-26 years) CV events are similar.

**Objective:** To use a combination of machine learning methods with deep phenotyping to differentiate 10-year versus lifetime predictors of CV outcomes.

**Design, Setting, and Participants:** This retrospective analysis used the prospectively collected data from the CARDIA (Coronary Artery Risk Development in Young Adults) study, a cohort of White and Black participants recruited from four clinical centers in the US. The analysis included 4314 participants, aged 23-35 years who were then followed up over 25 years through August 2018.

**Main Outcomes and Measures:** 449 variables collected in 1990-91 from imaging and noninvasive tests, questionnaires, and biomarker panels were included. We used machine learning techniques to identify the top-20 predictors of both 10-year and lifetime (10-26 years) CV events (coronary heart disease, myocardial infarction, acute coronary syndrome, stroke, transient ischemic attack, heart failure, peripheral arterial disease, and CV death).

**Results:** Kidney disease, family history of CV disease, and echocardiographic parameters of left ventricular systolic and diastolic dysfunction, and hypertrophy were important markers of 10-year CV events. Traditional risk factors and indices of body size featured heavily as top predictors of lifetime CV risk. Among the different machine learning techniques, Random Survival Forest and Nnet-survival performed the best (C-index of 0.80 for 10-year and 0.72 for lifetime). These models outperformed Cox models including traditional CV risk factors.

**Conclusions and Relevance:** Family history of CVD, kidney disease, and subclinical phenotyping of CVD using echocardiography are important for 10-year risk estimation. However, traditional CV risk factors alone may be adequate in estimating lifetime CV risk.

**Key Points:** *Question:* Do machine learning (ML) and deep learning (DL)-based survival analysis models help differentiate 10-year versus lifetime predictors of cardiovascular (CV) outcomes in young adults?

*Findings:* In this retrospective analysis of 4314 participants in the CARDIA study, ML and DL survival analysis improved CVD risk prediction over traditional Cox models and revealed the top 20 predictors among 449 variables. Top 10-year risk predictors include kidney disease, family history of CV disease, and echocardiographic parameters, where as traditional risk factors and indices of body size featured heavily as top predictors of lifetime CV risk.

*Meaning:* Family history, kidney disease, and subclinical phenotyping of CVD using echocardiography play a prominent role for 10-year risk estimation, while traditional CV risk factors alone may be adequate in estimating lifetime CV risk in young adults.

## Introduction

Epidemiological studies provide a framework for development of cardiovascular disease (CVD) risk prediction models, and have been central in deriving commonly used prediction algorithms such as the American College of Cardiology/American Heart Association Atherosclerotic Cardiovascular Disease (ACC/AHA ASCVD) risk score.^1^ Long-term studies like the Framingham, ARIC, MESA, CARDIA and several other studies have contributed substantially to our knowledge about the importance of lifestyle and environmental factors in CVD development.^2^ Prior studies have shown that while 82% of US adults are at low 10-year risk, two-thirds of this group, or 87 million people, are at high lifetime (>10 years) risk. Such a stepwise stratification system is thought to aid efforts in risk communication, and guide risk factor counseling and monitoring. Identification of those with high lifetime risk at a younger age may yield a greater benefit over lifecourse, if preventive strategies are implemented early in life.^3^ Identification of young adult populations at high risk of short-term cardiovascular (CV) events, and the factors associated with event onset remain important in the identification of optimal therapeutic strategies.

The use of traditional methods such as the Cox Proportional Hazard Models (Cox-PHM) in big datasets are limited because of the large number of existing covariates, correlation and non-linearity of different measures including potential complex interactions as well as the possibility of model over-fitting. Data mining methods, such as Random Survival Forests (RSF) using a non-parametric decision tree approach, and neural networks, allow us to overcome these potential limitations to a large extent while increasing the possibility of novel biomarker discover.^4^ To this extent, the main goal of our study was to identify the top predictors of 10-year and lifetime (> 10 years) risk. We used data from the (Coronary Artery Risk Development in Young Adults) CARDIA study, a large population-based epidemiologic study, which includes meticulously collected data on sociodemographic characteristics, risk factors, behaviors, imaging, and lifecourse exposures (over 25 years of follow-up). The overall goals of our study were to (a) compare the performance of machine learning (ML) and deeply phenotyping in comparison to Cox-PHM and traditional CV risk scores for CV event prediction; (b) identify the most important predictors of incident CVD for 10-year vs lifetime risk; and (c) understand the role of subclinical markers in 10-year vs. lifetime risk estimation.

## Methods

### Study Population and Candidate Variables

The design of the CARDIA study has been described elsewhere.^5^ Briefly, CARDIA is a prospective, observational cohort study of 5115 White and Black men and women aged 18 to 30 years, at four centers in the United States (Birmingham, AL.; Chicago, IL; Minneapolis, MN; and Oakland, CA). The cohort is approximately balanced regarding age, race, sex, and educational level. Participants have been followed since 1985, with regular exam visits scheduled every 2-5 years.^6^ Each exam has collected a wide variety of variables believed to be related to heart disease. The institutional review board of each participating institution approved the study protocol and all participants gave informed consent.

For this study, Exam-3 (Year-5) was defined as the baseline because it was the exam that contained the most phenotypic data in participants, before they reached 35 years of age. We used all variables available at baseline to predict the participants’ risk of CVD over 26 years of follow-up post-baseline. Among 4351 participants examined in Exam-3, 4314 participants had less than 25% missing data on candidate variables and were included. Candidate variables included questionnaires (dietary, physical exercise patterns, substance use), demographics, traditional risk factors, anthropometry, medication use, results of phlebotomy, urine chemistry, pulmonary function, psychological assessment, and echocardiography.^7–10^

### Outcome Definition

The CARDIA study events ascertainment protocol is described in detail in the eMethods in the Supplement. For this study, the first CVD event was used as the endpoint.^11,12^ and events were recorded through August 2018. The primary composite outcome was incident CVD, which included coronary heart disease (CHD, myocardial infarction, acute coronary syndrome, or CHD death, including fatal myocardial infarction), stroke, transient ischemic attack (TIA), hospitalization for heart failure, intervention for peripheral arterial disease, or death from CV causes. Participants who died from a non-cardiac cause were treated as censored cases in the survival models at time of death.

### Statistical Analysis

Figure 1 shows the schematic of the statistical analysis procedures. Data indexing and missing data handling followed pre-defined protocols (details in the Supplement, eFigure 1) and were performed when required. Random forest multiple imputation was used. The data were split into training-testing datasets with a 80:20 ratio. A total of 25 pairs of training-testing sets were made, in a 5×5-fold cross-validation fashion. Stratified splitting was used to ensure the same class ratio (non-CVD vs CVD) in the training sets and testing sets.. In short, the overall data resampling strategy followed a nested cross-validation scheme, where the inner loop is for hyperparameter tuning (details are provided in the Supplement), while the outer, 5×5-fold loop was used to estimate the generalized performance. The nested, repeated cross-validation strategy has been shown to be an optimal estimator of the true error^13,14^ and requires less computational time.^15^

**Figure 1:**
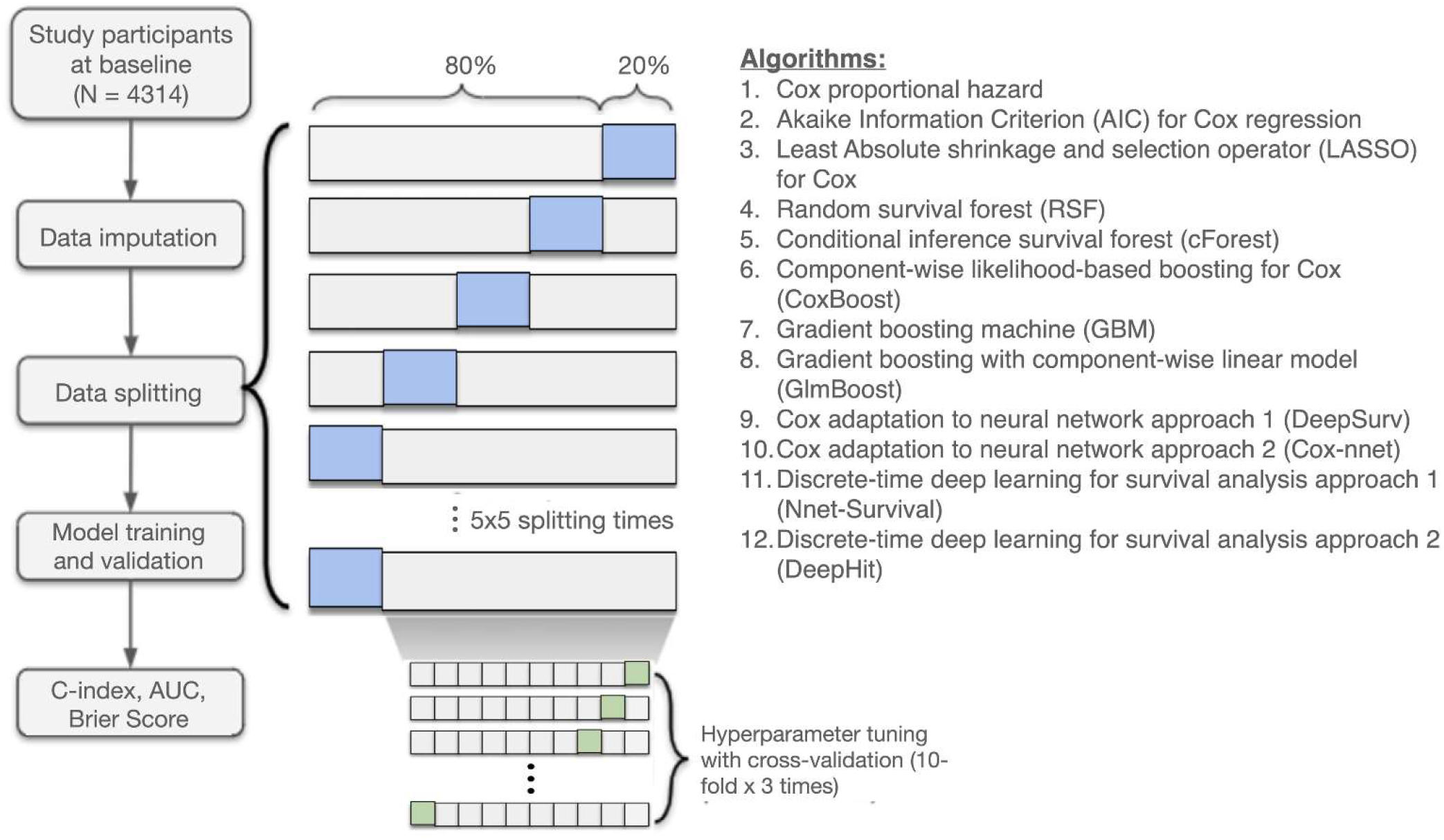
A flowchart describing the general framework of the study. Models were built using the training data sets (white), and the test data sets (blue) were used for computing the C-index, time-varying area under the curve (AUC), and the Brier score.

### Models Tested

Overall, we tested 12 algorithms, for 10-year and lifetime CVD event prediction, over three variable subsets: all variables, top 20 most important variables after using variable selection as detailed below, and nine traditional risk factors. This accounted for 72 models - 12 (algorithms) times 2 (10-year vs lifetime) times 3 (all variables, top-20 variables, and traditional risk factors). In addition, we also tested the ASCVD risk score for 10-year and lifetime risk estimation. The 12 algorithms (Figure 1) included three traditional statistical methods, five ML survival methods, and four deep learning (DL) survival methods. Details of the algorithms are described in the eMethods in the Supplement. We define 10-year prediction as predicting any CVD event within 0-10 years after the baseline exam, and included those with events or censored by 10 years after baseline. Those who experienced an event or were censored after 10 years were treated as censored at the 10-year mark. Lifetime event prediction used any incident CVD event between 10-26 years post-baseline. Hence, lifetime prediction excluded those with events or those that were censored before 10 years (conditioned on no events by 10 years). The origin for lifetime prediction was set at 10 years after post-baseline.

A total of 449 variables from CARDIA baseline (Exam-3) were used in the analysis. Variable importance was obtained the best performing model that used all variables, which was RSF. The measure of importance that we used is the mean of the minimal depth of the maximal subtree (highest point in the tree of a variable) over the entire forest, but we also looked at permutation testing (eTable 8). Essentially, in the minimum depth method, variables appearing higher on the tree have a higher rank. We used maximal subtree to rank variables because maximal subtree was shown to be better than permutation testing by both theory and simulation.^16^

### Performance Evaluation

The performance of each model was quantified using three metrics: time-dependent concordance index (C-index),^17^ cumulative time-dependent area under the receiver-operating curve (AUC) for discrimination,^18^ and Brier Score^19^ for calibration. We used the time-dependent C-index developed by Gerd’s et al^17^ which uses inverse probability of censoring weights when comparing the predicted and true ranks. The *t*-year cumulative AUC, is essentially the probability that a randomly chosen case has a higher predicted risk than a randomly chosen non-case at time *t*. The Brier Score measures the mean squared difference between the predicted probabilities and the actual outcomes. Higher C-index, higher AUC, and lower Brier Score indicate better prediction performance.

## Results

Among 4314 participants included in the analysis, the average age at baseline was 29 years with 55% women, 51.4% black and 48.6% white. At baseline, 1.6% of the participants had diabetes, 3.8% classified as hypertensive, and 36.5% were current or former smokers. Over a median of 25.87 years of follow-up post-baseline (interquartile range, 24.49– 25.12 years), 309 all-cause deaths, and 216 incident CVD events were identified (Table 1).

**Table 1.** Characteristics of the CARDIA participants at baseline (1990-91). Values inside parentheses denote standard deviation unless noted otherwise. The number of participants was 4314.

| Variable | Value |
| --- | --- |
| Age during Y5 Exam, year | 29.94 (3.37) |
| Sex (% female) | 45.3 |
| Race (% black) | 51.4 |
| Body mass index, kg/m<sup>2</sup> | 26.2 (5.5) |
| Diabetes mellitus (% diagnosed) | 1.6 |
| Systolic blood pressure, mmHg | 108.5 (12.9) |
| Diastolic blood pressure, mmHg | 66.0 (10.0) |
| Use of hypertensive medication (%) | 1.4 |
| Hypertension (%) | 3.8 |
| Age high blood pressure was diagnosed among hypertensive group | 24.3 (6.8) |
| Heart rate, bpm | 66.4 (9.5) |
| Smoking status |  |
| % current | 24.5 |
| % former | 12.1 |
| Smoking years | 9.9 (6.9) |
| Number of cigarettes smoked per day among current smokers | 13.0 (9.0) |
| Total cholesterol, mg/dL | 178.3 (31.4) |
| High-density lipoprotein cholesterol, mg/dL | 53.1 (13.0) |
| Low-density lipoprotein cholesterol, mg/dL | 108.5 (32.1) |
| Triglycerides, mg/dL | 80.8 (72.2) |
| Cholesterol-lowering medication (%) | 0.22 |
| Kidney problems (%) | 4.6 |
| Kidney stone (%) | 1.2 |
| Nephritis/ Glomerulonephritis (%) | 0.4 |
| LV Mass, grams | 161.03 (46.12) |
| M-mode: LV Post Wall Thickness – Diastole, cm | 0.85 (0.14) |
| M-mode: Left Atrial Dimension, cm | 3.52 (0.47) |
| M-mode: LV Expected Fractional Shortening, % | 27.54 (2.98) |
| M-mode: LVESS/LVFS Ratio, 10<sup>3</sup> dynes/cm<sup>2</sup>/% | 3.45 (2.15) |
| M-mode: LV Internal Dimension in Systole, cm | 3.19 (0.45) |
| M-mode: LV End Systolic Stress, 10<sup>3</sup> dynes/cm<sup>2</sup> | 116.38 (31.74) |
| DOP: 1st 3rd Diastolic Filling Fraction | 0.10 (0.028) |
| DOP: PFVA/PFVE Ratio | 0.59 (0.15) |
| DOP: Mitral Flow Velocity Integral - Early Diastole | 0.13 (0.03) |
| DOP: Mitral Peak Velocity - Late Diastole, m/s | 0.048 (0.015) |
| DOP: Pulmonary Artery Acceleration Time, ms | 139.67 (29.86) |
| Event by end of follow-up |  |
| CVD, n (%) | 216 (5.01) |
| All-cause death, n (%) | 309 (7.16) |

CVD: cardiovascular disease; LV, left ventricular; LVESS: LV end-systolic stress; LVFS: LV fractional shortening; DOP: Doppler echocardiography; PFVA/PFVE: Peak flow velocity A-to-E ratio.
\*Smoking regularly is defined as at least 5 cigs/week almost every week

### Comparison of Prediction Models

Figure 2 shows the performances of the top models in each variable subset over time. eTable 2 in the Supplement shows the performance (quantified by the C-index) of all 74 prediction models using three variable subsets, for 10-year and lifetime event prediction. Additional model performance metrics (e.g., AUC and Brier Score) are presented in the Supplement. The best models, characterized by the highest AUC, using all available variables were the RSF and Nnet-survival. Since RSF is also capable of providing variable importance, the top 20 predictors derived from RSF were fed into 12 algorithms to generate 24 top-20 parsimonious prediction models. RSF and Nnet-survival using the top 20 variables were again the best performing models both for 10-year (C-index=0.80), and lifetime CVD event prediction (C-index=0.72).

**Figure 2:**
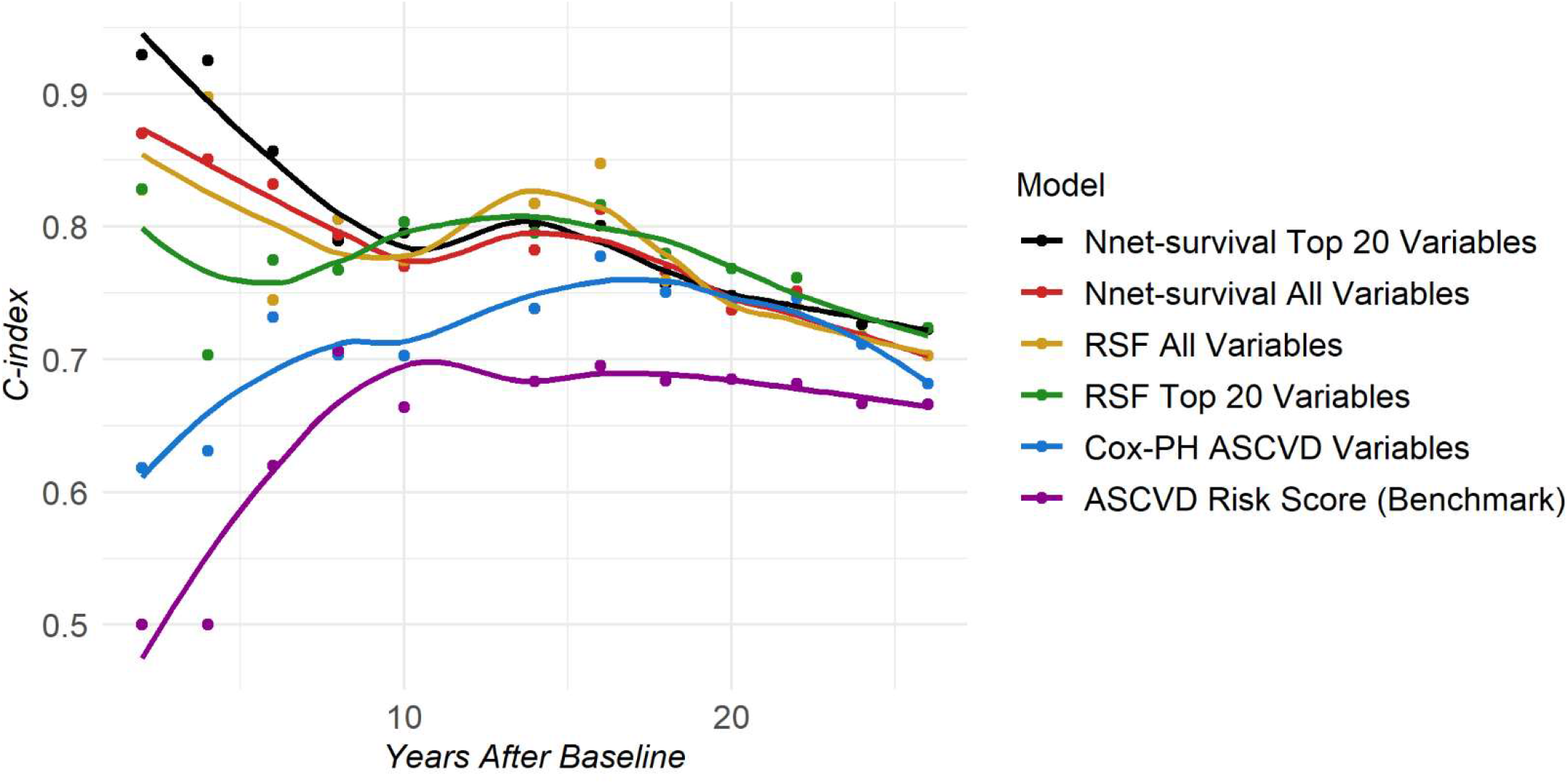
The median C-index over time for the baseline ACC/AHA ASCVD risk score model and the best performing models in each variable category: all variables, top 20 RSF-ranked variables, and ACC/AHA ASCVD variables only. RSF: Random Survival Forests; Cox-PH: Cox Proportional Hazards; ASCVD: atherosclerotic cardiovascular disease

For 10-year prediction, the difference in performance between models using 20 variables or more and models using only ASCVD variables was large, with as much as a 40% improved performance between years two to four and up to 13% at 10 years (Figure 2). The performance gap between ASCVD and non-ASCVD models decreased gradually over time. For lifetime prediction, the C-index of most ML models decreased 4% to 8% over 16 years, while the clinical reference model (ASCVD risk score) stayed at around 0.66-0.67 C-index.

Across variable subsets, models using top 20 variables performed the best. By using just the top 20 predictors and omitting noise from less important variables, the C-index increased up to 10% for 10-year prediction and up to 7% for lifetime prediction, compared to using all variables (eTable 2). The best ML models using ASCVD variables gave a C-index of 0.72 for 10-year prediction and 0.69 for lifetime prediction, while the C-index of the Cox model was 0.70 and 0.68, respectively. (eTable 2).

Of all algorithms, models trained by Nnet-survival and RSF were consistently the best among the different subsets (Figure 2, eTable 2).

### Predictors of Outcome

Figure 3 shows the ranking of all 449 variables grouped by category. The top 20 predictors for 10-year and lifetime CVD prediction are provided in Table 2. The top 20 predictors for CVD for all years is provided in the Supplement. eFigure 6 shows Lowess plots of survival probability over the range of values for the top-20 variables. All 449 variables were ranked using the ranking method from the best performing model to select the top predictors of CVD.

**Figure 3:**
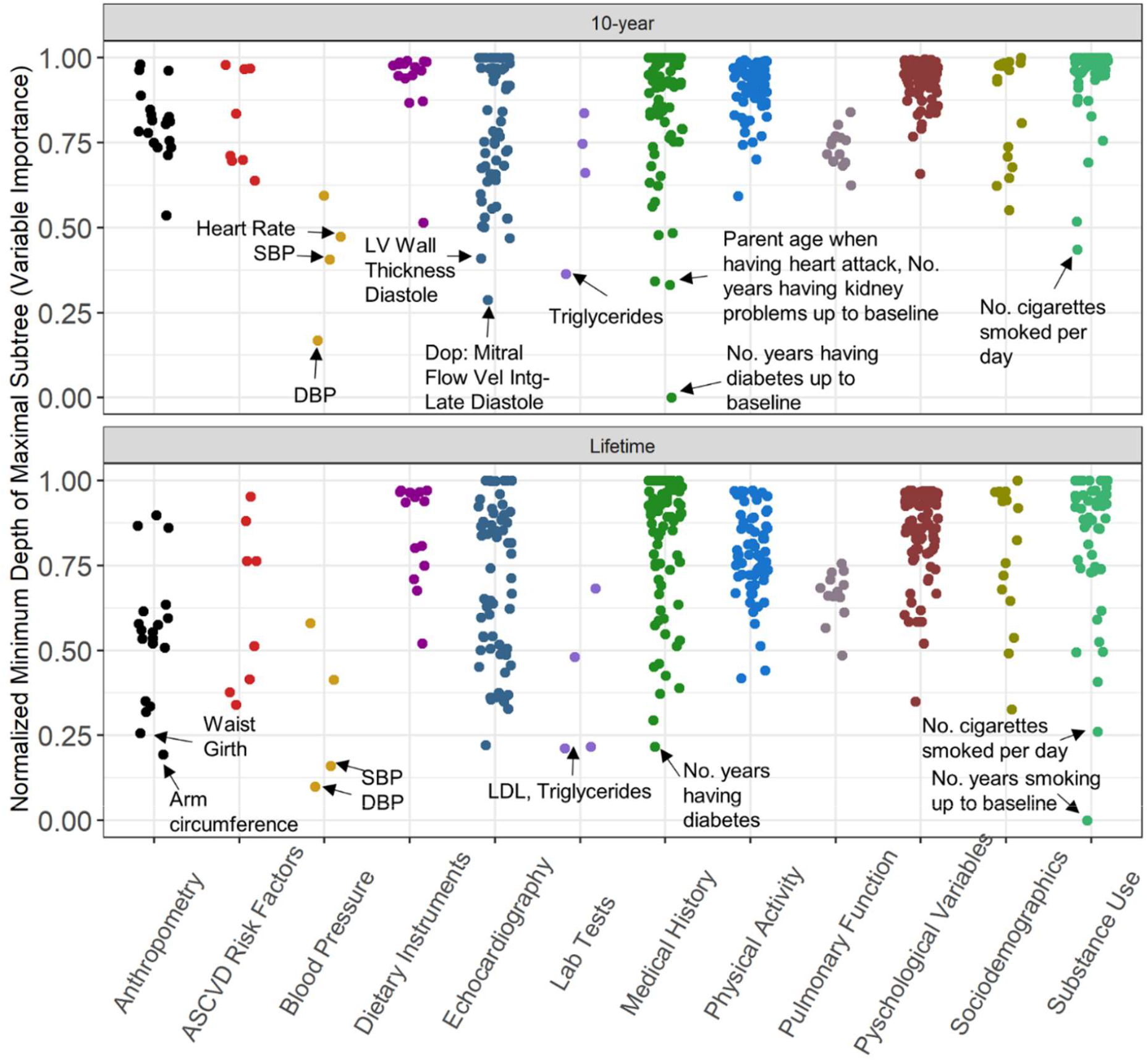
Dot plots showing the normalized variable importance for each of the 449 variables used in analysis in 10-year (top panel) and lifetime CVD prediction (bottom panel). The color of the dots represents the category of the variable. The variable importance is measured using the minimum depth of the maximal subtree in the Random Survival Forest model. The variable importance is normalized to range from 0 (most important) to 1 (least important). The top 10 most important variables are annotated.

**Table 2.** The top-20 ranked variables by the averaged variable importance from the Random Survival Forest method for the outcome of interest (CVD) using minimum depth of maximal subtree. RMD: relative minimal depth (0 is most important, 1 is least important).

| *Short term* |  |  | *Lifetime* |  |
| --- | --- | --- | --- | --- |
| **Rank** | **Variable** | **RMD** | **Variable** | **RMD** |
| 1 | # Years Having Diabetes by baseline Exam | 0.00 | How Many Years Smoked Regularly by baseline Exam | 0.00 |
| 2 | Average DBP | 0.17 | Average DBP | 0.10 |
| 3 | DOP: Mitral Flow Velocity Integral - Late Diastole | 0.29 | Average SBP | 0.16 |
| 4 | # Years Having Kidney Problems by baseline Exam | 0.33 | Arm Circumference | 0.19 |
| 5 | Parent's Age When Heart Attack Occurred | 0.34 | LDL Cholesterol | 0.21 |
| 6 | Triglycerides | 0.36 | Triglycerides | 0.22 |
| 7 | Average SBP | 0.41 | # Years Having Diabetes by baseline Exam | 0.22 |
| 8 | Heart Rate (Beats Per Minute) | 0.41 | DOP: AFVI/EFVI Ratio | 0.22 |
| 9 | Cigarettes, # Smoked/day | 0.44 | Waist Girth | 0.26 |
| 10 | M-mode: LV Post Wall Thickness – Diastole | 0.47 | Cigarettes, # Smoked/day | 0.26 |
| 11 | Pulse Pressure | 0.47 | # Years Having High Cholesterol by baseline Exam | 0.29 |
| 12 | # Years Having Nephritis/glomerulonephritis by baseline exam | 0.48 | Chest Girth | 0.32 |
| 13 | # Years Having Kidney Stone Diagnosed | 0.48 | Education Level: Grade of School Completed | 0.33 |
| 14 | Dop: Mitral Flow Velocity Integral - Early Diastole | 0.50 | DOP: PFVA/PFVE Ratio | 0.33 |
| 15 | M-mode: Left Atrial Dimension | 0.50 | Hip Girth | 0.34 |
| 16 | High blood pressure | 0.51 | Total HDL Cholesterol | 0.34 |
| 17 | How Many Years Smoked Regularly? | 0.52 | I Have Disturbing Thoughts | 0.35 |
| 18 | M-mode: LV Internal Dimension in Systole | 0.53 | M-mode: Left Atrial Dimension | 0.35 |
| 19 | M-mode: LVESS/LVFS Ratio | 0.53 | Subject's Weight | 0.35 |
| 20 | DOP: PFVA/PFVE Ratio | 0.53 | Body Surface Area | 0.36 |

The relative variable importance (RVI) of each variable can be assessed using the normalized minimal depth of the maximal subtree. The normalized RVI values vary from 0 (most important) to 1 (least important). CVD indicates cardiovascular disease; LV, left ventricular; DBP, Diastolic Blood Pressure; SBP: Systolic Blood Pressure; DOP: Doppler echocardiography; LVESS: LV end-systolic stress; LVFS: LV fractional shortening; DOP: Doppler echocardiography; PFVA/PFVE: Peak flow velocity A-to-E ratio; AFVI/EFVI: atrial to early diastolic flow velocity integral.
\*Smoking regularly is defined as at least 5 cigs/week almost every week

Duration of diabetes, diastolic blood pressure, systolic blood pressure, and triglycerides were consistently among the top 7 predictors of both 10-year and lifetime CVD events. Number of years smoking regularly, PFVA/PFVE Ratio (peak flow velocity A-peak to E-peak ratio), and left atrial dimension were among the top-20 incident CVD event predictors for both 10-year and lifetime (Table 2).

The importance of specific variables was different in short term and lifetime prediction models. For 10-year, the most important variable was duration of diabetes, while the next important variable was relatively less important (Figure 3, Table 2). In contrast, for lifetime risk, variables had similar relative importance. Smoking duration (defined as at least 5 cigarettes/week almost every week) and low-density lipoprotein (LDL) cholesterol were relatively more important for lifetime risk as opposed to 10-year risk (Figure 3). Anthropometry variables (waist girth, chest girth, arm circumference, and bodyweight) were more important for lifetime risk as opposed to 10-year (Figure 3). Psychological factors, such as measures of anxiety by questionnaire also rose to the top 20 lifetime CVD predictors.

Variables capturing kidney-related conditions and family history of heart attack were among the top 20 10-year predictors but not among the top lifetime predictors. Additionally, echocardiographic phenotypes reflecting left ventricular (LV) hypertrophy, reduced contractility, left atrial (LA) dilatation and diastolic dysfunction, were among the top 20 10-year predictors while only subclinical echocardiographic markers of diastolic dysfunction were among the top 20 lifetime predictors.

Regarding variable directionality, having diabetes for a longer time, longer smoking years, more cigarettes smoked per day, higher blood pressure, higher levels of triglycerides, cholesterol and LDL, as well as larger waist girth and greater body surface area were associated with higher chance of developing CVD (eFigure 6). Additionally, frequent anxiety and fewer education years were important CVD predictors. and lower Brier Score indicate better prediction performance.

## Discussion

Our results in a young population (23-35 years-old at baseline) suggest that ML/DL methods are well suited for meaningful risk prediction in extensively phenotyped large-scale epidemiological cohorts, particularly for 10-year CVD event prediction. RSF and Nnet-survival provided better prediction over standard risk scores and other ML and statistical techniques. An initial step of RSF-based variable selection followed by survival analysis also allowed for improved prediction, without problems of overfitting and nonconvergence while accounting for nonlinearities. Kidney pathologies were a non-traditional risk factor associated with 10-year risk, and echocardiographic parameters were strongly associated with 10-year risk. The results also confirm the importance of traditional risk factor tracking to monitor CVD risk in young populations, particularly over longer time periods across the lifespan. In this regard, obesity and related anthropometric measures were strongly associated with lifetime risk of CV events.

In our analysis, demographic variables of age, sex, and ethnicity were not among the top-20 predictors. The CARDIA population at baseline were young and between a narrow age-range of a little over a decade (23-35 years). This could have contributed to age not being among the top-20 markers. Age, sex, and ethnicity are associated with cardiac structure and function from echocardiography, which are more proximal to the disease itself, and thus may have been expressed through these factors in the top variables. In this regard, sex and race were not important predictors of CVD in this younger cohort although the findings were similar to our prior study using a ML approach in an older population.^4^ Conversely, as opposed to our prior study identifying markers of future CVD in older populations (MESA),^4^ in this analysis in the CARDIA population, well-known traditional risk factors remained among the top-20 important markers.^20,21^ These findings further highlight the particular importance of risk factor control and knowledge of CVD risk factors starting at a very young age.^22^ However, these discrepancies have to be considered in the context that not all biomarkers were measured in both the studies.

Premature CVD events (≤age 45) versus those occurring at middle age or later (> age 45), are thought to result from differences in the type and intensity of exposures to risk factors. Epidemiologically, CVD events increase exponentially from middle age on, but cost the most in relative individual and societal burden when occurring prematurely.^23,24^ Targeted screening strategies in the young may help identify risk factors that can isolate high-risk individuals.^25^ An important finding of the study is the differences in risk factors associated with lifetime and 10-year events. Parental history of heart disease was strongly associated with 10-year risk as has been observed in studies before.^26–28^ Kidney problems were associated with worse short-term prognosis. Prior studies in CARDIA have shown the association of kidney stones with subclinical atherosclerosis, and have also highlighted the importance of decreased kidney function in promoting greater CVD risk.^29–31^ LV mass and systolic function were important predictors of 10-year events.^32^ Indeed, a low CV risk profile in young adulthood is associated with more favorable LV mass, LV relative wall thickness, and LA size.^33,34^

In general, there were fewer subclinical echocardiographic features in the top predictors for lifetime risk. Indeed, prior work from the CARDIA study has shown a healthy lifestyle at young age is associated with a low CV profile at middle age.^35^ Obesity and abnormal body habitus were associated with worse lifetime prognosis. Obesity at a young age is associated with progressive worsening of structure and function over several years before being manifest as CVD events.^36^ Cumulative exposure to excessive adiposity is linked more strongly to incident CVD than BMI alone.^37^ LV diastolic dysfunction was the only subclinical echocardiographic marker predictive of lifetime risk. In a prior analysis to determine the prevalence and prognosis associated with diastolic dysfunction in CARDIA, the authors found that the prevalence of any diastolic abnormalities was 10.4%, and was associated with markedly increased risk for mortality and CV events, particularly after 10 years.^38^ Smoking status and years of smoking while also important for 10-year risk prediction, was the most important variable for lifetime risk prediction. Smoking is associated with subclinical cardiac dysfunction and an excess of minor and major ailments.^39–41^ CARDIA has also shown how high blood pressure and glycemic abnormalities at a young age are associated with a worse CV risk profile later in life.^42,43^

One of the goals of our study was to test a wide range of ML algorithms to identify the most robust ones. Each algorithm has its own advantages and disadvantages, and these are discussed extensively in the Supplement. In short, while traditional Cox models are easy to interpret, the underlying assumptions and lack of convergence for high-dimensional data are problematic. ML methods, which can be used both as a variable selection tool as well as a prediction algorithm, overcome these limitations and show improved performance. Newer DL techniques also provide improved prediction performance but at the cost of computational time. In our study, Nnet-survival from the DL techniques and RSF from the statistical ML class resulted in the best performing CVD prediction models.

### Limitations

Our study has several limitations. The data used at baseline was obtained in 1990, therefore general population characteristics may have changed over the last 30 years, and generalizability to the current youth has to be explored with caution. Second, external validation is challenging because cohorts of young adults with long-term follow-up that have extensive phenotyping similar to CARDIA are sparse.

## Conclusions

We show deep phenotyping in conjunction with ML improves CV event prediction over and above the use of traditional statistical approaches and risk factors among young adults for 10-year risk estimation. Family history of CVD, kidney disease, and subclinical phenotyping of CVD using echocardiography are important for 10-year risk estimation. However, traditional CV risk factors alone may be adequate in estimating lifetime CV risk.

## Data Availability

The CARDIA data can be requested via the study website https://www.cardia.dopm.uab.edu/. Data access is available through the CARDIA Coordinating Center following approval by the CARDIA Publications and Presentations Committee. Please see the study website for further details: http://www.cardia.dopm.uab.edu/invitation-to-new-investigators.

## Acknowledgements

The Coronary Artery Risk Development in Young Adults Study (CARDIA) is conducted and supported by the National Heart, Lung, and Blood Institute (NHLBI) in collaboration with the University of Alabama at Birmingham (HHSN268201800005I & HHSN268201800007I), Northwestern University (HHSN268201800003I), University of Minnesota (HHSN268201800006I), and Kaiser Foundation Research Institute (HHSN268201800004I). This manuscript has been reviewed by CARDIA for scientific content. The views expressed in this manuscript are those of the authors and do not necessarily represent the views of the National Heart, Lung, and Blood Institute; the National Institutes of Health; or the U.S. Department of Health and Human Services.

## References

1. Goff Jr DC, Lloyd-Jones DM, Bennett G, et al. 2013 ACC/AHA guideline on the assessment of cardiovascular risk: a report of the American College of Cardiology/American Heart Association Task Force on Practice Guidelines. Circulation. 2014;129(25_suppl_2):S49–S73.

2. Lloyd-Jones DM. Cardiovascular risk prediction: basic concepts, current status, and future directions. Circulation. 2010;121(15):1768–1777.

3. Arnett DK, Blumenthal RS, Albert MA, et al. 2019 ACC/AHA guideline on the primary prevention of cardiovascular disease: a report of the American College of Cardiology/American Heart Association Task Force on Clinical Practice Guidelines. Circulation. 2019;140(11):e596–e646.

4. Ambale-Venkatesh B, Yang X, Wu CO, et al. Cardiovascular event prediction by machine learning: the multi-ethnic study of atherosclerosis. Circ Res. 2017;121(9):1092–1101.

5. Friedman GD, Cutter GR, Donahue RP, et al. Cardia: study design, recruitment, and some characteristics of the examined subjects. J Clin Epidemiol. 1988;41(11):1105–1116. doi:10.1016/0895-4356(88)90080-7

6. Loria CM, Liu K, Lewis CE, et al. Early Adult Risk Factor Levels and Subsequent Coronary Artery Calcification. The CARDIA Study. J Am Coll Cardiol. Published online 2007. doi:10.1016/j.jacc.2007.03.009

7. Gardin JM, Wagenknecht LE, Anton-Culver H, et al. Relationship of cardiovascular risk factors to echocardiographic left ventricular mass in healthy young black and white adult men and women: the CARDIA study. Circulation. 1995;92(3):380–387.

8. Burke GL, Bild DE, Hilner JE, Folsom AR, Wagenknecht LE, Sidney S. Differences in weight gain in relation to race, gender, age and education in young adults: the CARDIA study. Ethn Health. 1996;1(4):327–335.

9. Kiefe CI, Williams OD, Greenlund KJ, Ulene V, Gardin JM, Raczynski JM. Health care access and seven-year change in cigarette smoking: The CARDIA study. Am J Prev Med. 1998;15(2):146–154.

10. Sidney S, Jacobs Jr DR, Haskell WL, et al. Comparison of two methods of assessing physical activity in the Coronary Artery Risk Development in Young Adults (CARDIA) Study. Am J Epidemiol. 1991;133(12):1231–1245.

11. Bibbins-Domingo K, Pletcher MJ, Lin F, et al. Racial Differences in Incident Heart Failure among Young Adults. New England Journal of Medicine. Published online 2009. doi:10.1056/nejmoa0807265

12. Armstrong AC, Jacobs DR, Gidding SS, et al. Framingham score and LV mass predict events in young adults: CARDIA study. Int J Cardiol. Published online 2014. doi:10.1016/j.ijcard.2014.01.003

13. Varma S, Simon R. Bias in error estimation when using cross-validation for model selection. BMC Bioinformatics. Published online 2006. doi:10.1186/1471-2105-7-91

14. Krstajic D, Buturovic LJ, Leahy DE, Thomas S. Cross-validation pitfalls when selecting and assessing regression and classification models. J Cheminform. Published online 2014. doi:10.1186/1758-2946-6-10

15. Kim JH. Estimating classification error rate: Repeated cross-validation, repeated hold-out and bootstrap. Comput Stat Data Anal. Published online 2009. doi:10.1016/j.csda.2009.04.009

16. Ishwaran H, Kogalur UB, Gorodeski EZ, Minn AJ, Lauer MS. High-dimensional variable selection for survival data. J Am Stat Assoc. Published online 2010. doi:10.1198/jasa.2009.tm08622

17. Gerds TA, Kattan MW, Schumacher M, Yu C. Estimating a time-dependent concordance index for survival prediction models with covariate dependent censoring. Stat Med. 2013;32(13):2173–2184.

18. Liang CJ, Heagerty PJ. A risk-based measure of time-varying prognostic discrimination for survival models. Biometrics. 2017;73(3):725–734.

19. Steyerberg EW, Vickers AJ, Cook NR, et al. Assessing the performance of prediction models: a framework for some traditional and novel measures. Epidemiology. 2010;21(1):128.

20. Liu K, Colangelo LA, Daviglus ML, et al. Can antihypertensive treatment restore the risk of cardiovascular disease to ideal levels? The Coronary Artery Risk Development in Young Adults (CARDIA) Study and the Multi-Ethnic Study of Atherosclerosis (MESA). J Am Heart Assoc. 2015;4(9):e002275.

21. Liu K, Wilkins JT, Colangelo LA, Lloyd-Jones DM. Does Lowering Low-Density Lipoprotein Cholesterol With Statin Restore Low Risk in Middle-Aged Adults? Analysis of the Observational MESA Study. J Am Heart Assoc. 2021;10(11):e019695.

22. Lynch EB, Liu K, Kiefe CI, Greenland P. Cardiovascular disease risk factor knowledge in young adults and 10-year change in risk factors: the Coronary Artery Risk Development in Young Adults (CARDIA) Study. Am J Epidemiol. 2006;164(12):1171–1179.

23. Sniderman AD, Thanassoulis G, Williams K, Pencina M. Risk of premature cardiovascular disease vs the number of premature cardiovascular events. JAMA Cardiol. 2016;1(4):492–494.

24. Kotseva K, Gerlier L, Sidelnikov E, et al. Patient and caregiver productivity loss and indirect costs associated with cardiovascular events in Europe. Eur J Prev Cardiol. 2019;26(11):1150–1157.

25. Lawson KD, Fenwick EAL, Pell ACH, Pell JP. Comparison of mass and targeted screening strategies for cardiovascular risk: simulation of the effectiveness, cost-effectiveness and coverage using a cross-sectional survey of 3921 people. Heart. 2010;96(3):208–212.

26. Parikh NI, Hwang SJ, Larson MG, et al. Parental occurrence of premature cardiovascular disease predicts increased coronary artery and abdominal aortic calcification in the Framingham Offspring and Third Generation cohorts. Circulation. 2007;116(13):1473–1481.

27. Chow CK, Islam S, Bautista L, et al. Parental history and myocardial infarction risk across the world: the INTERHEART Study. J Am Coll Cardiol. 2011;57(5):619–627.

28. Burke GL, Savage PJ, Sprafka JM, et al. Relation of risk factor levels in young adulthood to parental history of disease. The CARDIA study. Circulation. 1991;84(3):1176–1187.

29. Peralta CA, Lin F, Shlipak MG, et al. Race differences in prevalence of chronic kidney disease among young adults using creatinine-based glomerular filtration rate-estimating equations. Nephrology Dialysis Transplantation. 2010;25(12):3934–3939.

30. Peralta CA, Vittinghoff E, Bansal N, et al. Trajectories of kidney function decline in young black and white adults with preserved GFR: results from the Coronary Artery Risk Development in Young Adults (CARDIA) study. American journal of kidney diseases. 2013;62(2):261–266.

31. Reiner AP, Kahn A, Eisner BH, et al. Kidney stones and subclinical atherosclerosis in young adults: the CARDIA study. J Urol. 2011;185(3):920–925.

32. Armstrong AC, Gidding S, Gjesdal O, Wu C, Bluemke DA, Lima JAC. LV mass assessed by echocardiography and CMR, cardiovascular outcomes, and medical practice. JACC Cardiovasc Imaging. 2012;5(8):837–848.

33. Gidding SS, Liu K, Colangelo LA, et al. Longitudinal determinants of left ventricular mass and geometry: the Coronary Artery Risk Development in Young Adults (CARDIA) Study. Circ Cardiovasc Imaging. 2013;6(5):769–775.

34. Gidding SS, Carnethon MR, Daniels S, et al. Low cardiovascular risk is associated with favorable left ventricular mass, left ventricular relative wall thickness, and left atrial size: the CARDIA study. Journal of the American Society of Echocardiography. 2010;23(8):816–822.

35. Liu K, Daviglus ML, Loria CM, et al. Healthy lifestyle through young adulthood and the presence of low cardiovascular disease risk profile in middle age: the Coronary Artery Risk Development in (Young) Adults (CARDIA) study. Circulation. 2012;125(8):996–1004.

36. Murthy VL, Abbasi SA, Siddique J, et al. Transitions in metabolic risk and long-term cardiovascular health: coronary artery risk development in young adults (CARDIA) Study. J Am Heart Assoc. 2016;5(10):e003934.

37. Reis JP, Allen N, Gunderson EP, et al. Excess body mass index-and waist circumference-years and incident cardiovascular disease: the CARDIA study. Obesity. 2015;23(4):879–885.

38. Desai CS, Colangelo LA, Liu K, et al. Prevalence, prospective risk markers, and prognosis associated with the presence of left ventricular diastolic dysfunction in young adults: the coronary artery risk development in young adults study. Am J Epidemiol. 2013;177(1):20–32.

39. Moreira HT, Armstrong AC, Nwabuo CC, et al. Association of smoking and right ventricular function in middle age: CARDIA study. Open Heart. 2020;7(1):e001270.

40. Gidding SS, Xie X, Liu K, Manolio T, Flack JM, Gardin JM. Cardiac function in smokers and nonsmokers: the CARDIA study. J Am Coll Cardiol. 1995;26(1):211–216.

41. Hozawa A, Houston T, Steffes MW, et al. The association of cigarette smoking with self-reported disease before middle age: the Coronary Artery Risk Development in Young Adults (CARDIA) study. Prev Med (Baltim). 2006;42(3):193–199.

42. Kishi S, Gidding SS, Reis JP, et al. Association of insulin resistance and glycemic metabolic abnormalities with LV structure and function in middle age: the CARDIA study. JACC Cardiovasc Imaging. 2017;10(2):105–114.

43. Allen NB, Siddique J, Wilkins JT, et al. Blood pressure trajectories in early adulthood and subclinical atherosclerosis in middle age. JAMA. 2014;311(5):490–497.

