## Supplemental Materials for "Lifetime vs 10-year Cardiovascular Disease Prediction in Young Adults Using Statistical Machine Learning and Deep Learning: The CARDIA Study"


**eMethods**

**eReferences**

**eTable 1.** List of algorithms employed for prediction of CVD in the CARDIA study

**eTable 2.** Comparison of predictive performance of all models used based on their C-index to predict CVD events.

**eTable 3.** Predictive Performance for Models Trained on All Variables, for 10-year and Lifetime Prediction.

**eTable 4.** Predictive Performance for Models Trained on ASCVD Variables, for 10-year and Lifetime Prediction.

**eTable 5.** Predictive Performance for Models Trained on Top 20 Ranked Variables, for 10-year and Lifetime Prediction

**eTable 6.** Predictive Performance for RSF Model Trained on All Variables, for CVD Prediction for All Follow-up Years After Y5 Exam

**eTable 7.** The Top-20 Ranked Variables by the Averaged Variable Importance from the Random Survival Forest Method for the Outcome of Interest (CVD) using Minimum Depth of Maximal Subtree, for all the Years (0-26).

**eTable 8.** The Top-20 Ranked Variables from the Random Survival Forest Method Using Permutation Test for the Outcome of Interest (CVD), for Lifetime Prediction

**eTable 9.** Deep learning final hyperparameters

**eFigure 1.** The pie chart shows the distribution of available data

**eFigure 2.** CVD Cumulative Incidence Function in the CARDIA cohort after Y5 Exam (Exam 3).

**eFigure 3.** Effect of number of trees on the out-of-bag error rate in random survival forest (top panel: 10-year prediction, bottom panel: lifetime prediction)

**eFigure 4.** Nested RSF For the Top Predictors of CVD by Year 10

**eFigure 5.** Nested RSF For the Top Predictors of CVD by Year 26

**eFigure 6.** Plots showing Lowess curves of the marginal effect on survival probability of the top-20 predictors for lifetime CVD outcome prediction (Year 10 to Year 26 after the Baseline exam).

**eMethods. CARDIA event ascertainment**

During their scheduled study examinations and yearly telephone interviews, each participant or designated proxy was asked about interim hospital admissions, outpatient procedures, and deaths. Medical records were requested for participants who had been hospitalized or received an outpatient revascularization procedure. Vital status was assessed every 6 months; medical and other death records were requested after consent had been obtained from the next of kin. Two physician members of the Endpoints Committee independently reviewed medical records and recorded information to adjudicate each possible CV or cerebrovascular event or underlying cause of death using specific definitions and a detailed manual of operations. If disagreement occurred between the primary reviewers, the case was reviewed by the full committee.

**eMethods. Algorithm introduction and comparison**

A total of 12 algorithms were used in the prediction models, with three traditional statistical methods, five machine learning survival methods, and four deep learning survival methods. The full list is shown in eTable 1.

Cox PH is frequently used in survival analysis studies because it is easy to understand and requires small computational time. However, Cox and even AIC-Cox don't converge for a large number of covariates, as shown in Table 2. LASSO-Cox mitigates the convergence problem and is often used for variable selection. However, it still suffers from assumptions about proportional hazards (the effect of each predictor variable is the same at all values of follow-up time) and about covariate linear contribution to the logarithm of the hazards ratio when in fact, the relationship could be quite complex and not linear.(1) The statistical machine learning class mitigates these problems, as they model survival times with greater flexibility without any distributional assumption.

Ensembled tree-based methods such as RSF and cForest are also known for being fully parametric, robust, and have been shown to achieve performance gains in a variety of medical cohorts.(2–4) Drawbacks of RSFs include bias towards favoring variables with many split points.(5) CForest, utilizing conditional inference, mitigates this selection bias by having a separate algorithm for selecting the best covariate to split on from the best split point search process.(6)

Unlike the forest methods which compose of many small trees, boosting methods combine weak and simple models sequentially to build a strong model. CoxBoost is an iterative "gradient boosting" method modified from the Cox-PH model. In CoxBoost, parameters are separated into individual partitions, and the partition that leads to the largest improvement in the penalized partial log likelihood is selected for that iteration.(7) Despite not being popularly used in survival analysis, boosting algorithms have also been shown to achieve high performance gains in classification tasks. However, boosting is sometimes prone to overfitting. The last algorithm category, deep learning-based survival methods are relatively new. The superior flexibility from the neural networks allows them to achieve very high-performance gain in theory.(8–11) DeepSurv and Cox-nnet are neural network adaptation of Cox PH, while the other two algorithms, Nnet-survival and DeepHit, do not assume proportional hazards and use discrete-time instead. They require much more computational time and complexity for hyperparameter tuning and have limited interpretability capability. Overall, Nnet-survival from the DL algorithm class and RSF from the statistical ML class resulted in the best performing CVD prediction models.

**eReferences**

1. Samuelsen SO. Exact Inference in the Proportional Hazard Model: Possibilities and Limitations. Lifetime Data Anal. 2003;9(3):239–60.

2. Gorodeski EZ, Ishwaran H, Kogalur UB, Blackstone EH, Hsich E, Zhang Z ming, et al. Use of Hundreds of Electrocardiographic Biomarkers for Prediction of Mortality in Postmenopausal Women. Circulation Cardiovasc Qual Outcomes. 2011;4(5):521–32.

3. Ishwaran H, Kogalur UB, Blackstone EH, Lauer MS. Random survival forests. Ann Appl Statistics. 2008;2(3):841–60.

4. Nasejje JB, Mwambi H, Dheda K, Lesosky M. A comparison of the conditional inference survival forest model to random survival forests based on a simulation study as well as on two applications with time-to-event data. Bmc Med Res Methodol. 2017;17(1):115.

5. Strobl C, Boulesteix AL, Zeileis A, Hothorn T. Bias in random forest variable importance measures: Illustrations, sources and a solution. Bmc Bioinformatics. 2007;8(1):25.

6. Wright MN, Dankowski T, Ziegler A. Unbiased split variable selection for random survival forests using maximally selected rank statistics. Stat Med. 2017;36(8):1272–84.

7. Bin RD. Boosting in Cox regression: a comparison between the likelihood-based and the model-based approaches with focus on the R-packages CoxBoost and mboost. Computation Stat. 2016;31(2):513–31.

8. Lee C, Yoon J, Schaar M van der. Dynamic-DeepHit: A Deep Learning Approach for Dynamic Survival Analysis With Competing Risks Based on Longitudinal Data. Ieee T Bio-med Eng. 2018;67(1):122–33.

9. Katzman JL, Shaham U, Cloninger A, Bates J, Jiang T, Kluger Y. DeepSurv: personalized treatment recommender system using a Cox proportional hazards deep neural network. Bmc Med Res Methodol. 2018;18(1):24.

10. Gensheimer MF, Narasimhan B. A scalable discrete-time survival model for neural networks. Peerj. 2019;7:e6257.

11. Ching T, Zhu X, Garmire LX. Cox-nnet: An artificial neural network method for prognosis prediction of high-throughput omics data. Plos Comput Biol. 2018;14(4):e1006076.

**eTable 1.** List of algorithms employed for prediction of CVD in the CARDIA study

| Name | Description | Package or GitHub name (in R, asterisk if in Python) |
|---|---|---|
| **Coxph** | Cox proportional hazard | Survival |
| **AIC-Cox** | Akaike Information Criterion for Cox regression | MASS |
| **LASSO-Cox** | Least absolute shrinkage and selection operator (L1) for Cox regression | Glmnet |
| **RSF** | Random survival forest | Rsfrc |
| **Cforest** | Conditional inference survival forest | Party |
| **CoxBoost** | Component-wise likelihood-based boosting for Cox | CoxBoost |
| **Gbm** | Gradient boosting machine | Gbm |
| **Glmboost** | Gradient boosting with component-wise linear model | Mboost |
| **Cox-nnet** | Cox proportional hazard adaptation to neural network approach 1 | *Cox-nnet[1] |
| **DeepSurv** | Cox proportional hazard adaptation to neural network approach 2 | *DeepSurv[2] |
| **Nnet-Survival** | Discrete-time deep learning approach for survival analysis approach 1 | *Nnet-survival[3] |
| **DeepHit** | Discrete-time deep learning approach for survival analysis approach 2 | *DeepHit[4] |

[1] http://garmiregroup.org/cox-nnet/docs/
[2] https://github.com/jaredleekatzman/DeepSurv
[3] https://github.com/MGensheimer/nnet-survival
[4] https://github.com/chl8856/DeepHit

**eTable 2.** Comparison of predictive performance of all models used based on their C-index to predict CVD events.

| *Algorithm* | *449 Variables* | | *Top 20 Variables* | | *9 Traditional Risk Factors* | |
|---|---|---|---|---|---|---|
| | **Short term** | **Lifetime** | **Short term** | **Lifetime** | **Short term** | **Lifetime** |
| ***ASCVD risk score*** | X | X | X | X | 0.67 (0.62, 0.72) | 0.66 (0.65, 0.68) |
| ***RSF*** | 0.76 (0.73, 0.79) | 0.70 (0.69, 0.72 | 0.80 (0.77, 0.83) | 0.72 (0.71, 0.74) | 0.70 (0.68, 0.73) | 0.67 (0.64, 0.7) |
| ***CForest*** | 0.76 (0.73, 0.79) | 0.71 (0.69, 0.74) | 0.80 (0.76, 0.84) | 0.72 (0.71, 0.74) | 0.72 (0.7, 0.75) | 0.66 (0.64, 0.69) |
| ***Cox*** | No convergence | No convergence | 0.70 (0.66, 0.75) | 0.72 (0.7, 0.73) | 0.70 (0.67, 0.74) | 0.68 (0.66, 0.71) |
| ***AIC-Cox*** | No convergence | No convergence | 0.73 (0.69, 0.76) | 0.70 (0.69, 0.72) | 0.68 (0.64, 0.72) | 0.68 (0.65, 0.71) |
| ***LASSO-Cox*** | 0.68 (0.64, 0.73 | 0.63 (060, 0.66) | 0.73 (0.69, 0.76) | 0.70 (0.69, 0.72) | 0.68 (0.64, 0.72) | 0.68 (0.65, 0.71) |
| ***CoxBoost*** | 0.63 (0.59, 0.66) | 0.70 (0.68, 0.72) | 0.76 (0.73, 0.79) | 0.71 (0.7, 0.72) | 0.69 (0.66, 0.73) | 0.64 (0.6, 0.69) |
| ***GBM*** | 0.67 (0.63, 0.72) | 0.68 (0.66, 0.70) | 0.78 (0.75, 0.81) | 0.72 (0.7, 0.73) | 0.60 (0.57, 0.64) | 0.65 (0.61, 0.68) |
| ***Glmboost*** | 0.63 (0.59, 0.67) | 0.70 (0.68, 0.73) | 0.73 (0.70, 0.76) | 0.71 (0.69, 0.72) | 0.63 (0.6, 0.67) | 0.63 (0.6, 0.65) |
| ***Nnet-survival*** | 0.77 (0.74, 0.80) | 0.70 (0.69, 0.72) | 0.80 (0.76, 0.83) | 0.72 (0.71, 0.73) | 0.71 (0.68, 0.74) | 0.66 (0.64, 0.68) |
| ***DeepSurv*** | 0.63 (0.59, 0.67) | 0.62 (0.60, 0.63) | 0.67 (0.61, 0.73) | 0.69 (0.68, 0.70) | 0.68 (0.65, 0.71) | 0.65 (0.63, 0.68) |
| ***Cox-nnet*** | 0.64 (0.62, 0.67) | 0.71 (0.68, 0.71) | 0.74 (0.71, 0.77) | 0.70 (0.68, 0.72) | 0.53 (0.51, 0.55) | 0.65 (0.64, 0.66) |
| ***DeepHit*** | 0.63 (0.59, 0.67) | 0.65 (0.62, 0.68) | 0.72 (0.66, 0.77) | 0.71 (0.69, 0.72) | 0.65 (0.62, 0.68) | 0.69 (0.67, 0.71) |

*Predictive performance in terms of C-index at endpoint for all 74 models using three variable subsets: all 449 variables, top 20 variables, and 9 traditional risk factors used in the ASCVD risk score equations (clinical baseline). 10-year prediction is defined as predicting CVD from 0 to 10 years after CARDIA Y5 exam (41 events out of 4314 at-risk participants, or 0.95% prevalence). Lifetime prediction is defined as prediction of CVD from 10 to 26 years after baseline (175 events out of 4200 at-risk participants, or 4.2% prevalence). Values inside parentheses denote the bootstrapped empirical 95% confident interval. Values outside parentheses denote the mean.*

*RSF: random survival forests; GBM: gradient boosting machines; ASCVD: atherosclerotic cardiovascular disease; AIC: Akaike information criteria.*

**eTable 3.** Predictive Performance for Models Trained on All Variables, for 10-year and Lifetime Prediction. 10-year prediction is defined as Predicting CVD from 0 To 10 Years After Cardia Y5 Exam (41 Events Out Of 4314 At-risk Participants, Or 0.95% Prevalence). Lifetime Prediction is defined as Predicting CVD from 10 To 26 Years After Cardia Y5 Exam (175 Events Out Of 4200 At-risk Participants, Or 4.2% Prevalence). Values Inside Parentheses Denote the Bootstrapped Empirical 95% Confident Interval. Values Outside Parentheses Denote the Mean.

| | | *10-year* | | | *Lifetime* | | |
|---|---|---|---|---|---|---|---|
| *Algorithm* | No. of variables | C-index | AUC | Brier | C-index | AUC | Brier |
| *ASCVD risk score* | 9 | 0.67 (0.62, 0.72) | 0.67 (0.62, 0.72) | 0.006 (0.005, 0.006) | 0.66 (0.65, 0.68) | 0.66 (0.65, 0.68) | 0.036 (0.035,0.038) |
| *RSF* | 449 | 0.76 (0.73, 0.79) | 0.76 (0.73, 0.79) | 0.006 (0.005, 0.006) | 0.70 (0.69, 0.72) | 0.70 (0.69, 0.72) | 0.036 (0.035,0.038) |
| *CForest* | 449 | 0.76 (0.73, 0.79) | 0.76 (0.74, 0.79) | 0.004 (0.005, 0.006) | 0.71 (0.69, 0.74) | 0.72 (0.68, 0.76) | 0.036 (0.035,0.037) |
| *Cox* | 449 | Did not converge | | | Did not converge | | |
| *AIC-Cox* | 449 | Did not converge | | | Did not converge | | |
| *LASSO-Cox* | 24 (21, 26) | 0.68 (0.64, 0.73) | 0.68 (0.64, 0.73) | 0.006 (0.005, 0.006) | 0.63 (060, 0.66) | 0.62 (0.57, 0.67) | 0.036 (0.035,0.037) |
| *CoxBoost* | 449 | 0.63 (0.59, 0.66) | 0.62 (0.59, 0.66) | 0.006 (0.005, 0.006) | 0.70 (0.68, 0.72) | 0.71 (0.67, 0.75) | 0.036 (0.035,0.037) |
| *Gbm* | 449 | 0.67 (0.63, 0.72) | 0.68 (0.63, 0.72) | 0.006 (0.005, 0.006) | 0.68 (0.66, 0.70) | 0.65 (0.62, 0.68) | 0.036 (0.035,0.037) |
| *Glmboost* | 449 | 0.63 (0.59, 0.67) | 0.63 (0.59, 0.68) | 0.005 (0.005, 0.006) | 0.70 (0.68, 0.73) | 0.72 (0.68, 0.73) | 0.035 (0.034, 0.036) |
| *Nnet-survival* | 449 | 0.77 (0.74, 0.80) | 0.77 (0.72, 0.82) | 0.006 (0.005, 0.006) | 0.70 (0.69, 0.72) | 0.70 (0.69, 0.72) | 0.035 (0.034, 0.036) |
| *DeepSurv* | 449 | 0.63 (0.59, 0.67) | 0.63 (0.59, 0.67) | 0.005 (0.005, 0.006) | 0.62 (0.60, 0.63) | 0.62 (0.60, 0.63) | 0.044 (0.043, 0.045) |
| *Cox-nnet* | 449 | 0.64 (0.62, 0.67) | 0.65 (0.62, 0.67) | 0.005 (0.005, 0.006) | 0.71 (0.68, 0.74) | 0.71 (0.67, 0.75) | 0.035 (0.034, 0.036) |
| *DeepHit* | 449 | 0.63 (0.59, 0.67) | 0.63 (0.59, 0.67) | 0.006 (0.005, 0.007) | 0.65 (0.62, 0.68) | 0.65 (0.62, 0.68) | 0.035 (0.034, 0.036) |

**eTable 4.** Predictive Performance for Models Trained on ASCVD Variables, for 10-year and Lifetime Prediction. 10-year prediction is defined as Predicting CVD from 0 To 10 Years After Cardia Y5 Exam (41 Events Out Of 4314 At-risk Participants, Or 0.95% Prevalence). Lifetime Prediction is defined as Predicting CVD from 10 To 26 Years After Cardia Y5 Exam (175 Events Out Of 4200 At-risk Participants, Or 4.2% Prevalence). Values Inside Parentheses Denote the Bootstrapped Empirical 95% Confident Interval. Values Outside Parentheses Denote the Mean.

| Model | No. of variables | 10-year | | | Lifetime | | |
|---|---|---|---|---|---|---|---|
| | | C-index | AUC | Brier | C-index | AUC | Brier |
| ASCVD risk score | 9 | 0.67 (0.62, 0.72) | 0.67 (0.62, 0.72) | 0.006 (0.005, 0.006) | 0.66 (0.65, 0.68) | 0.66 (0.65, 0.68) | 0.036 (0.035,0.038) |
| RSF | 9 | 0.7 (0.68, 0.73) | 0.7 (0.68, 0.73) | 0.005 (0.005, 0.006) | 0.67 (0.64, 0.7) | 0.64 (0.59, 0.7) | 0.036 (0.035,0.038) |
| CForest | 9 | 0.72 (0.70, 0.75) | 0.73 (0.7, 0.75) | 0.006 (0.005, 0.006) | 0.66 (0.64, 0.69) | 0.64 (0.58, 0.7) | 0.036 (0.035,0.038) |
| Cox | 9 | 0.70 (0.67, 0.74) | 0.70 (0.67, 0.74) | 0.006 (0.005, 0.006) | 0.68 (0.66, 0.71) | 0.67 (0.61, 0.72) | 0.036 (0.035,0.038) |
| AIC-Cox | 9 | 0.68 (0.64, 0.72) | 0.68 (0.65, 0.72) | 0.006 (0.005, 0.006) | 0.68 (0.65, 0.71) | 0.68 (0.65, 0.71) | 0.036 (0.035,0.038) |
| LASSO-Cox | 9 | 0.68 (0.64, 0.72) | 0.68 (0.65, 0.72) | 0.006 (0.005, 0.006) | 0.68 (0.65, 0.71) | 0.68 (0.65, 0.71) | 0.036 (0.035,0.038) |
| CoxBoost | 9 | 0.69 (0.66, 0.73) | 0.70 (0.66, 0.73) | 0.005 (0.005, 0.006) | 0.64 (0.6, 0.69) | 0.63 (0.57, 0.69) | 0.036 (0.035,0.038) |
| Gbm | 9 | 0.60 (0.57, 0.64) | 0.60 (0.57, 0.64) | 0.006 (0.005, 0.006) | 0.65 (0.61, 0.68) | 0.61 (0.56, 0.66) | 0.052 (0.049,0.056) |
| Glmboost | 9 | 0.63 (0.6, 0.67) | 0.64 (0.6, 0.67) | 0.006 (0.005, 0.006) | 0.63 (0.6, 0.65) | 0.59 (0.53, 0.64) | 0.036 (0.034,0.037) |
| Nnet-survival | 9 | 0.71 (0.68, 0.74) | 0.71 (0.68, 0.75) | 0.006 (0.005, 0.006) | 0.66 (0.64, 0.68) | 0.64 (0.59, 0.69) | 0.042 (0.042, 0.043) |
| DeepSurv | 9 | 0.68 (0.65, 0.71) | 0.68 (0.65, 0.72) | 0.005 (0.005, 0.006) | 0.65 (0.63, 0.68) | 0.60 (0.54, 0.65) | 0.036 (0.034, 0.037) |
| Cox-nnet | 9 | 0.53 (0.51, 0.55) | 0.53 (0.51, 0.55) | 0.005 (0.005, 0.006) | 0.65 (0.64, 0.66) | 0.65 (0.64, 0.66) | 0.044 (0.044, 0.045) |
| DeepHit | 9 | 0.65 (0.62, 0.68) | 0.65 (0.62, 0.68) | 0.006 (0.005, 0.006) | 0.69 (0.67, 0.71) | 0.69 (0.67, 0.71) | 0.11 (0.08, 0.14) |

**eTable 5.** Predictive Performance for Models Trained on Top 20 Ranked Variables, for 10-year and Lifetime Prediction. 10-year prediction is defined as Predicting CVD from 0 To 10 Years After Cardia Y5 Exam (41 Events Out Of 4314 At-risk Participants, Or 0.95% Prevalence). Lifetime Prediction is defined as Predicting CVD from 10 To 26 Years After Cardia Y5 Exam (175 Events Out Of 4200 At-risk Participants, Or 4.2% Prevalence). Values Inside Parentheses Denote the Bootstrapped Empirical 95% Confident Interval. Values Outside Parentheses Denote the Mean.

| Model | No. of variables | 10-year | | | Lifetime | | |
|---|---|---|---|---|---|---|---|
| | | C-index | AUC | Brier | C-index | AUC | Brier |
| RSF | 20 | 0.80 (0.77, 0.83) | 0.80 (0.77, 0.83) | 0.006 (0.005, 0.006) | 0.72 (0.71, 0.74) | 0.72 (0.71, 0.74) | 0.036 (0.035,0.038) |
| CForest | 20 | 0.80 (0.76, 0.84) | 0.80 (0.77, 0.84) | 0.006 (0.005, 0.006) | 0.72 (0.71, 0.74) | 0.72 (0.68, 0.76) | 0.036 (0.035,0.036) |
| Cox | 20 | 0.70 (0.66, 0.75) | 0.71 (0.66, 0.75) | 0.006 (0.005, 0.006) | 0.72 (0.7, 0.73) | 0.72 (0.68, 0.77) | 0.036 (0.035,0.036) |
| AIC-Cox | 14 | 0.73 (0.69, 0.76) | 0.73 (0.69, 0.76) | 0.006 (0.005, 0.006) | 0.70 (0.69, 0.72) | 0.71 (0.66, 0.75) | 0.036 (0.035,0.036) |
| LASSO-Cox | 15 (15, 16) | 0.73 (0.69, 0.76) | 0.73 (0.69, 0.76) | 0.006 (0.005, 0.006) | 0.70 (0.69, 0.72) | 0.71 (0.66, 0.75) | 0.036 (0.035,0.036) |
| CoxBoost | 20 | 0.76 (0.73, 0.79) | 0.76 (0.73, 0.79) | 0.006 (0.005, 0.006) | 0.71 (0.7, 0.72) | 0.71 (0.66, 0.75) | 0.035 (0.035,0.036) |
| Gbm | 20 | 0.78 (0.75, 0.81) | 0.78 (0.75, 0.82) | 0.006 (0.005, 0.006) | 0.72 (0.7, 0.73) | 0.66 (0.62, 0.70) | 0.044 (0.041, 0.047) |
| Glmboost | 20 | 0.73 (0.7, 0.76) | 0.73 (0.7, 0.76) | 0.005 (0.005, 0.006) | 0.71 (0.69, 0.72) | 0.70 (0.66, 0.75) | 0.035 (0.035,0.036) |
| Nnet-survival | 20 | 0.80 (0.76, 0.83) | 0.80 (0.76, 0.83) | 0.005 (0.005, 0.006) | 0.72 (0.71, 0.73) | 0.72 (0.67, 0.76) | 0.036 (0.035,0.038) |
| DeepSurv | 20 | 0.67 (0.61, 0.73) | 0.67 (0.61, 0.73) | 0.005 (0.005, 0.006) | 0.69 (0.68, 0.70) | 0.69 (0.65, 0.73) | 0.036 (0.033, 0.037) |
| Cox-nnet | 20 | 0.74 (0.71, 0.77) | 0.74 (0.71, 0.77) | 0.005 (0.005, 0.006) | 0.70 (0.68, 0.71) | 0.68 (0.60, 0.77) | 0.036 (0.033, 0.037) |
| DeepHit | 20 | 0.72 (0.66, 0.77) | 0.72 (0.66, 0.77) | 0.097 (0.086, 0.105) | 0.71 (0.69, 0.72) | 0.71 (0.69, 0.72) | 0.036 (0.033, 0.037) |

**eTable 6.** Predictive Performance for RSF Model Trained on All Variables, for CVD Prediction for All Follow-up Years After Y5 Exam (216 Events Out Of 4314 At-risk Participants, Or 5.01% Prevalence). Performances at 10 Years and 26 Years After y5 Exam are shown. Values Inside Parentheses Denote the Bootstrapped Empirical 95% Confident Interval. Values Outside Parentheses Denote the Mean.

| | | *10 Years After Y5 Exam* | | | *26 Years After Y5 Exam* | | |
|---|---|---|---|---|---|---|---|
| *Algorithm* | No. of variables | C-index | AUC | Brier | C-index | AUC | Brier |
| *RSF* | 449 | 0.79 (0.75, 0.82) | 0.79 (0.75, 0.82) | 0.003 (0.002, 0.003) | 0.74 (0.73, 0.76) | 0.72 (0.70, 0.74) | 0.043 (0.042, 0.044) |

**eTable 7.** The Top-20 Ranked Variables by the Averaged Variable Importance from the Random Survival Forest Method for the Outcome of Interest (CVD) using Minimum Depth of Maximal Subtree, for all the Years (0-26). RMD: Relative Minimal Depth (0 is Most important, 1 is Least Important).

| **Rank** | **Variable Description** | **RVI** |
|---|---|---|
| 1 | # Years Having Diabetes by Y5 Exam | 0.00 |
| 2 | # Years Smoked Regularly by Y5 Exam | 0.02 |
| 3 | Average DBP | 0.07 |
| 4 | Average SBP | 0.19 |
| 5 | Triglycerides (Mg/dl) | 0.22 |
| 6 | Arm Circumference in Cm | 0.24 |
| 7 | LDL Cholesterol (Mg/dl) | 0.24 |
| 8 | Cigarettes, # Smoked/day | 0.27 |
| 9 | Dop: AFVI/EFVI Ratio | 0.29 |
| 10 | Dop: Mitral Flow Vel Intg - Late Dia | 0.30 |
| 11 | Parent's Age When Heart Attack Occurred | 0.32 |
| 12 | M-mode: Left Atrial Dimension | 0.33 |
| 13 | M-mode: LV Post Wall Thickness - Diastole | 0.34 |
| 14 | Grade of School Completed | 0.34 |
| 15 | Heart Rate (Beats Per Minute) | 0.35 |
| 16 | Waist Girth (Cm) | 0.36 |
| 17 | M-mode: LV End Systolic Stress | 0.36 |
| 18 | # Years Having Kidney Stone by Y5 Exam | 0.38 |
| 19 | Dop: PFVA/PFVE Ratio | 0.38 |
| 20 | Dop: Mitral Peak Vel - Late Diastole | 0.38 |

*The relative variable importance (RVI) of each variable can be assessed using the normalized minimal depth of the maximal subtree. The normalized RVI values vary from 0 (most important) to 1 (least important). CVD indicates cardiovascular disease; LV, left ventricular; DBP, Diastolic Blood Pressure; SBP: Systolic Blood Pressure; DOP: Doppler echocardiography; LVESS: LV end-systolic stress; LVFS: LV fractional shortening; DOP: Doppler echocardiography; PFVA/PFVE: Peak flow velocity A-to-E ratio; AFVI/EFVI: atrial to early diastolic flow velocity integral.*

**Smoking regularly is defined as at least 5 cigs/week almost every week*

**eTable 8.** The Top-20 Ranked Variables from the Random Survival Forest Method Using Permutation Test for the Outcome of Interest (CVD), for Lifetime Prediction (Predicting CVD from 10 To 26 Years After Cardia Y5 Exam). RVI: Relative Variable Importance (1 is most important, 0 is least important).

| **Rank** | **Variable Description** | **RVI** |
|---|---|---|
| 1 | # Years Having Diabetes | 1.00 |
| 2 | How Many Yrs. Smoked Regularly? | 0.91 |
| 3 | Elevated Blood Pressure | 0.84 |
| 4 | M-mode: Lv Expected Frac Shortening | 0.82 |
| 5 | M-mode: Lv End Systolic Stress | 0.80 |
| 6 | Triglycerides (Mg/dl) | 0.69 |
| 7 | Waist Girth (Cm) | 0.69 |
| 8 | Average Dbp (Sum_d/2) | 0.68 |
| 9 | Hdl Cholesterol (Mg/dl) | 0.66 |
| 10 | Ovaries Been Removed | 0.55 |
| 11 | M-mode: Left Atrial Dimension | 0.53 |
| 12 | M-mode: Left Vent. Mass (Coc Calculated) | 0.52 |
| 13 | Exam Center | 0.46 |
| 14 | Average Sbp (Sum_s/2) | 0.45 |
| 15 | Dop: Afvi/efvi Ratio | 0.43 |
| 16 | Total Cholesterol (Mg/dl) | 0.39 |
| 17 | Ldl Cholesterol (Mg/dl) | 0.36 |
| 18 | Cigarettes, # Smoked/day | 0.36 |
| 19 | M-mode: Lvess/lvfs Ratio | 0.35 |
| 20 | I Have Disturbing Thoughts | 0.35 |

**eTable 9.** Deep learning final hyperparameters

Models with all variables

| Hyperparameter | Nnet-survival | DeepSurv | Cox-nnet | DeepHit |
|---|---|---|---|---|
| Gradient Descent | Adam | Adam | Adam | Adam |
| Learning rate | 1.601e-4 | 1.117e-4 | 0.0001 | 0.1 |
| Learning rate decay | 0.001 | 0.1213 | 0.9 | 0 |
| L2 Regularization | 0.4894 | 0.6596 | 0.1 | 0 |
| L1 Regularization | 0 | 3.574e-2 | 0 | 0 |
| Dropout | NA | 0.158 | 0 | 0.265 |
| Batch size | 256 | NA | 0 | 256 |
| Number epochs | 1000 | 462 | 200,000 | 1000 |
| Patience (for early stopping) | 22 | 112 | 1000 | 200 |
| # Dense layers | 2 | 2 | 1 | 2 |
| # Nodes in layer 1 | 3 | 392 | 143 | 32 |
| # Nodes in layer 2 | 52 | 21 | NA | 32 |
| # Discrete time intervals | 52 | NA | NA | 52 |
| Momentum | 0.85 | 0.9 | 0.9 | NA |
| Alpha | NA | NA | NA | 0.1572 |
| Sigma | NA | NA | NA | 0.4556 |

Models with the top 20 variables

| Hyperparameter | Nnet-survival | DeepSurv | Cox-nnet | DeepHit |
|---|---|---|---|---|
| Gradient Descent | Adam | Nesterov | Adam | Adam |
| Learning rate | 0.001 | 0.0001 | 0.0001 | 0.1 |
| Learning rate decay | 0.001 | 0.001 | 0.9 | NA |
| L2 Regularization | 0.1 | 0.001 | 1e-6 | NA |
| L1 Regularization | 0 | 0 | 0 | 0 |
| Dropout | NA | 0.4000 | 0 | 0.100 |
| Batch size | 256 | NA | 0 | 256 |
| Number epochs | 1000 | 10 | 200,000 | 10,000 |
| Patience (for early stopping) | 20 | 112 | 1000 | 200 |
| # Dense layers | 2 | 1 | 1 | 2 |
| # Nodes in layer 1 | 4 | 7 | 143 | 32 |
| # Nodes in layer 2 | 52 | NA | NA | 32 |
| # Discrete time intervals | 52 | NA | NA | 52 |
| Momentum | 0.9 | 0.9 | 0.9 | NA |
| Alpha | NA | NA | NA | 0.2 |
| Sigma | NA | NA | NA | 0.1 |

Models with ASCVD variables

| Hyperparameter | Nnet-survival | DeepSurv | Cox-nnet | DeepHit |
|---|---|---|---|---|
| Gradient Descent | Adam | Adam | Adam | Adam |
| Learning rate | 0.001 | 1.114e-4 | 0.0001 | 0.001 |
| Learning rate decay | 0.001 | 0.1213 | 0.9 | NA |
| L2 Regularization | 0.1 | 0.1455 | 0.1 | NA |
| L1 Regularization | 0 | 0.3333 | 0 | 0 |
| Dropout | NA | 0.158 | 0 | 0.265 |
| Batch size | 256 | NA | 0 | 256 |
| Number epochs | 50,000 | 954 | 200,000 | 1000 |
| Patience (for early stopping) | 22 | 1111 | 1000 | 200 |
| # Dense layers | 2 | 2 | 1 | 2 |
| # Nodes in layer 1 | 4 | 31 | 143 | 9 |
| # Nodes in layer 2 | 52 | 33 | NA | 9 |
| # Discrete time intervals | 52 | NA | NA | 52 |
| Momentum | 0.9 | 0.8685 | 0.9 | NA |
| Alpha | NA | NA | NA | 0.2397 |
| Sigma | NA | NA | NA | 0.3205 |

**eFigure 1.** The pie chart shows the distribution of available data. The legends indicate the percentage of participants that the variables were available in. Of all 500+ variables considered, 3% of the variables were available in less than 50% of the population, while 90.4% of the variables were available in more than 90% of the population. Variables with less than 50% availability were removed from the analysis. After removing duplicated variables, 449 variables were the final number of variables that were included in the modeling phase. Missing values for these 449 variables were filled using the random forest imputation method *rf_impute()* in the 'RandomforestSRC' package.

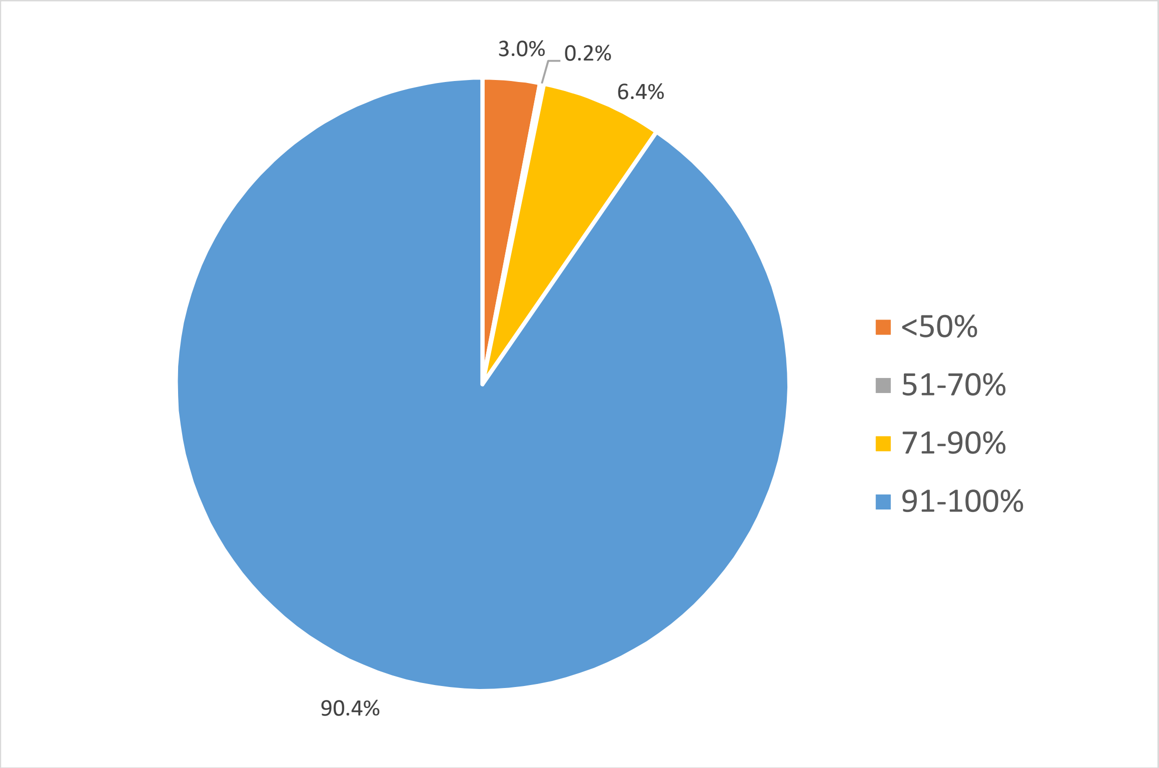

**eFigure 2.** CVD Cumulative Incidence Function in the CARDIA cohort after Y5 Exam (Exam 3).

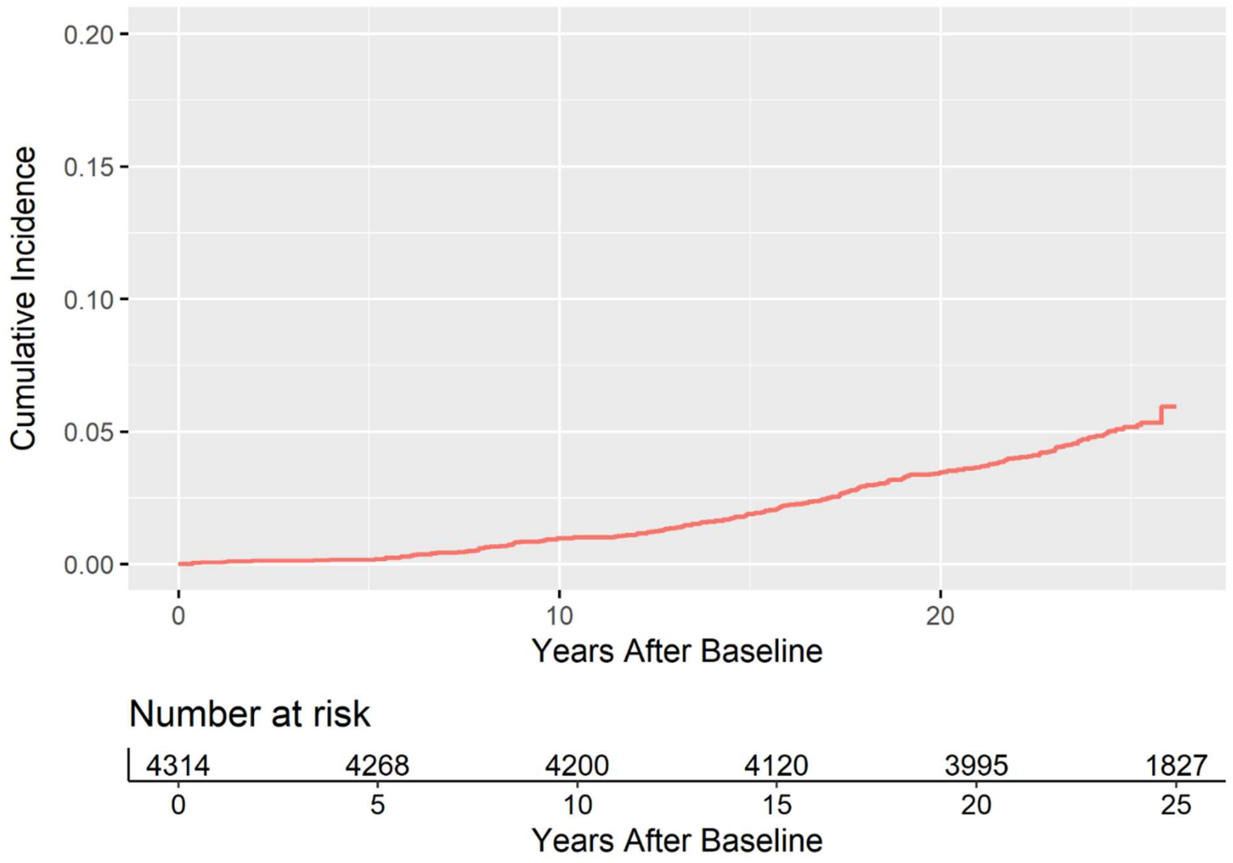

**eFigure 3.** Effect of number of trees on the out-of-bag error rate in random survival forest (top panel: 10-year prediction, bottom panel: lifetime prediction).

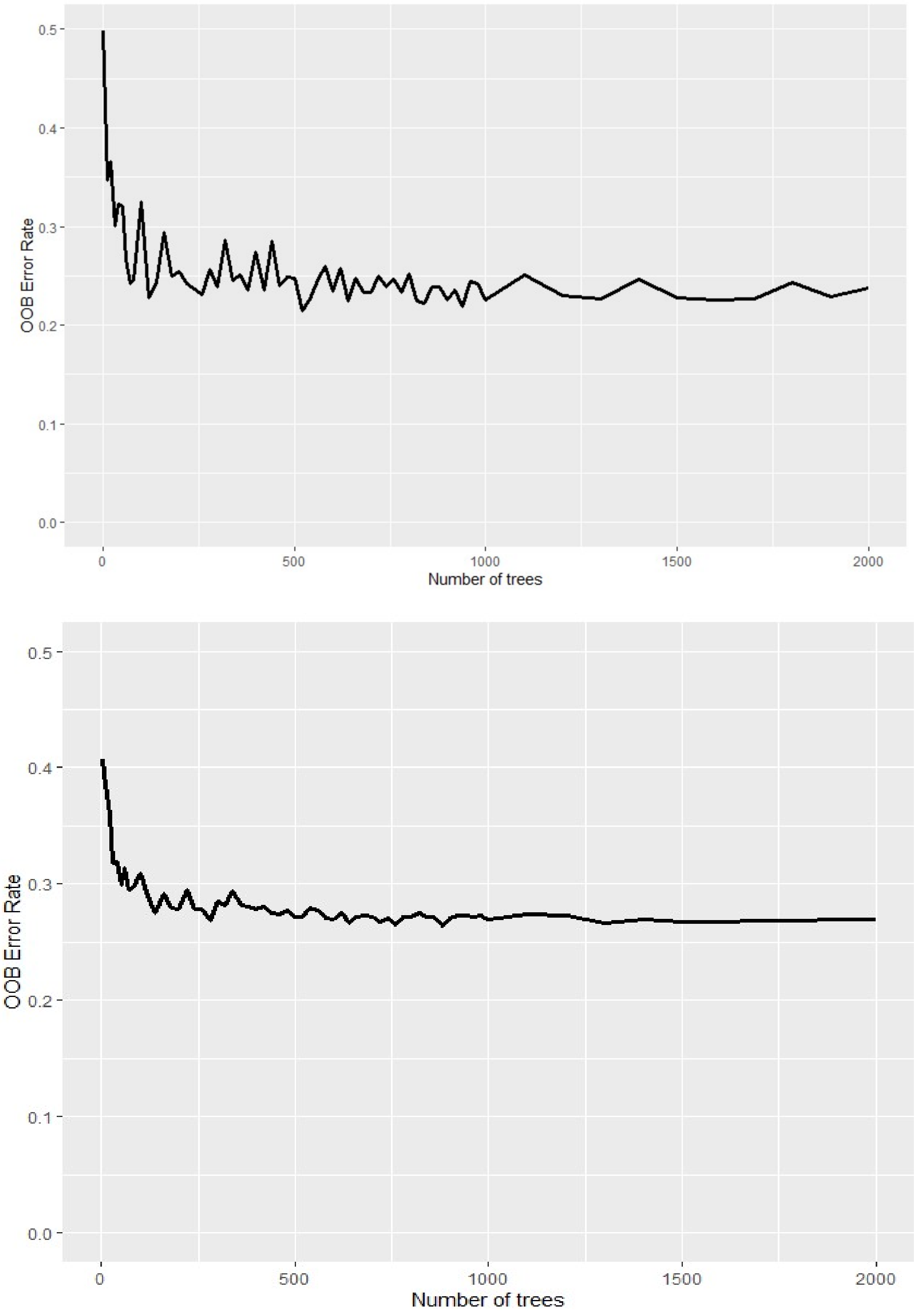

**eFigure 4.** Nested RSF For the Top Predictors of CVD by Year 10 (zoomed version on top).

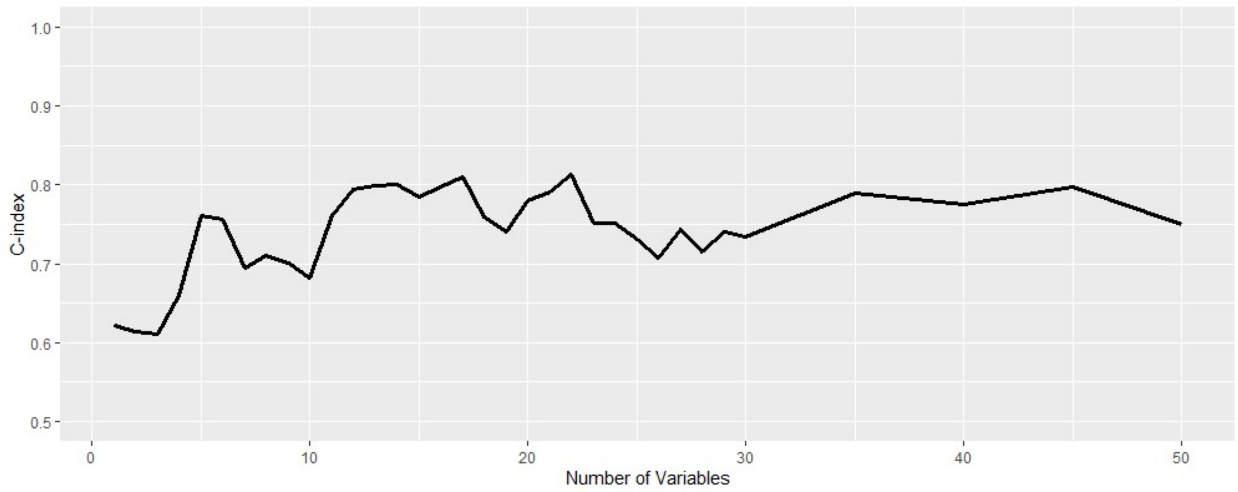


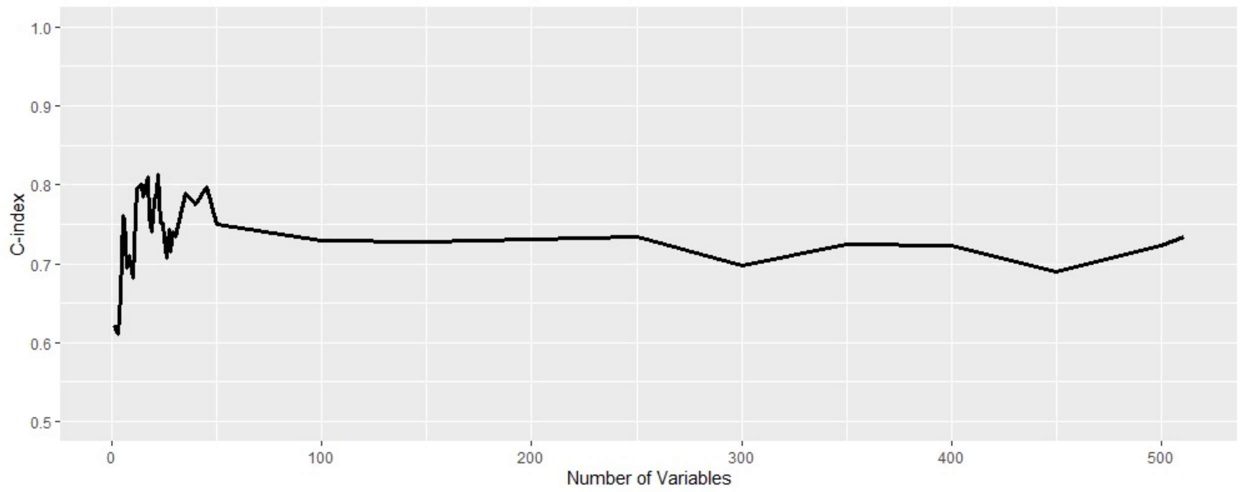

**eFigure 5.** Nested RSF For the Top Predictors of CVD by Year 26 (zoomed version on top).

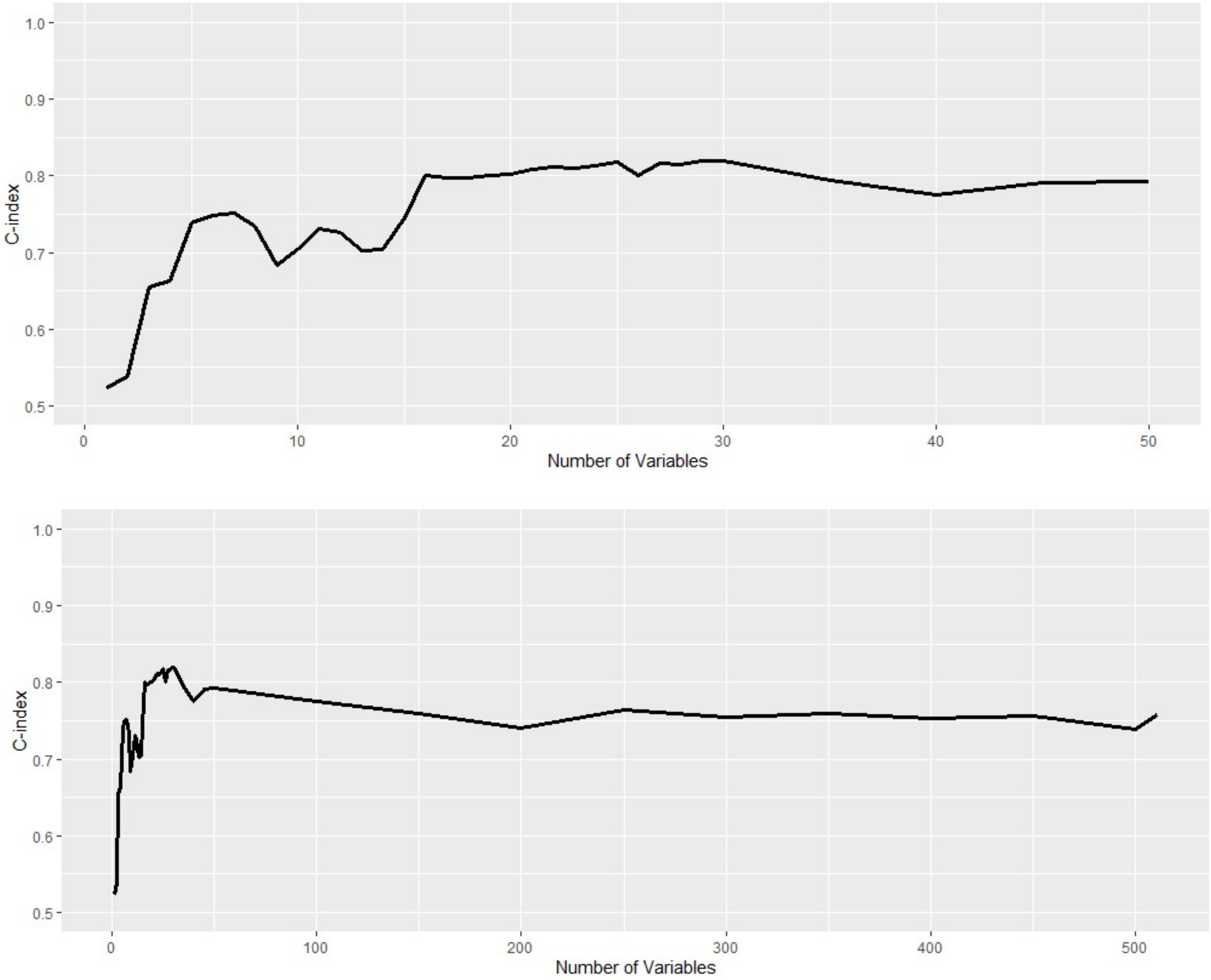

**eFigure 6.** Plots showing Lowess curves of the marginal effect on survival probability of the top-20 predictors for lifetime CVD outcome prediction (Year 10 to Year 26 after the Baseline exam). All the 'number of years' variables were up to the baseline exam.

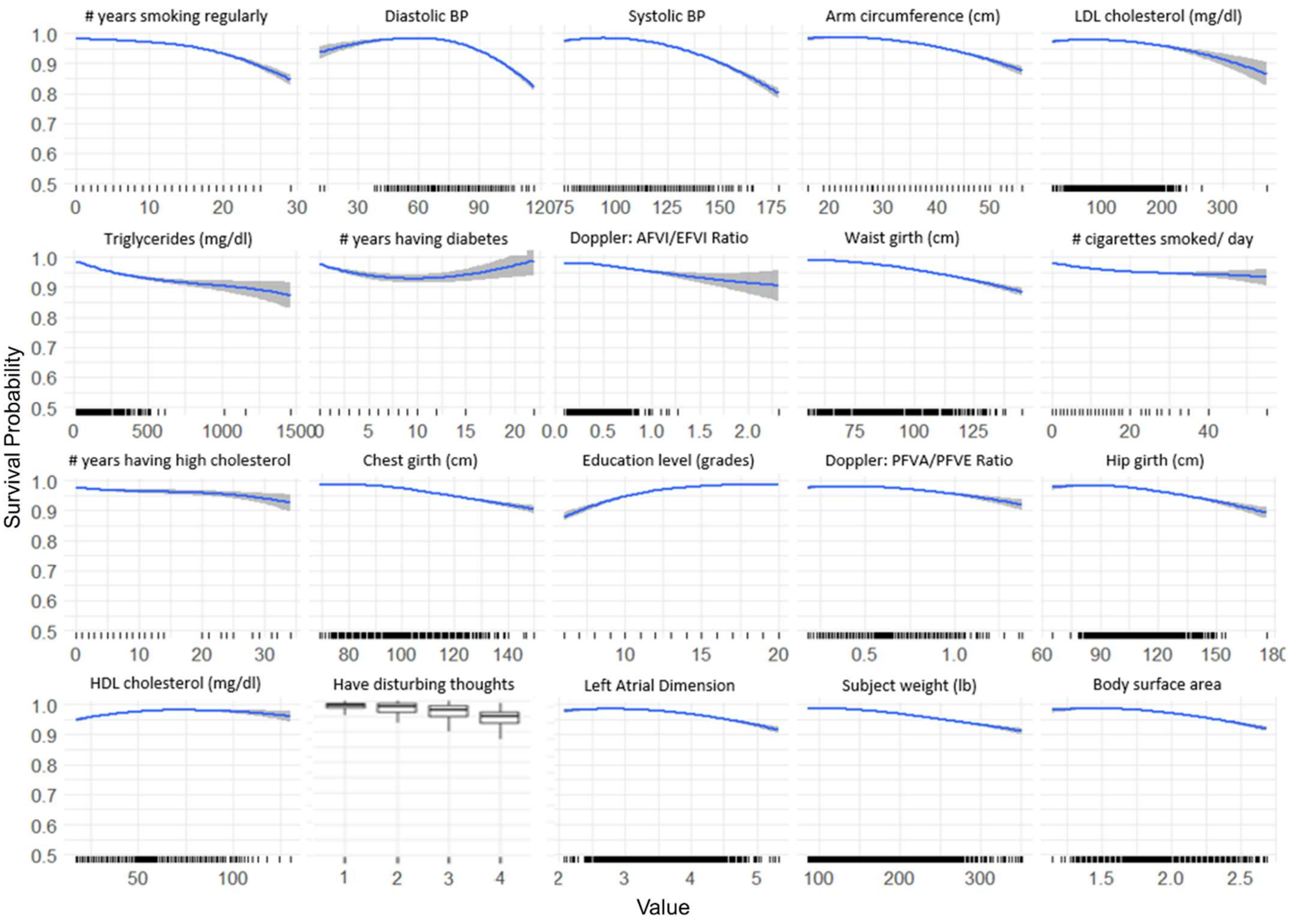
